# *KCNN3* genetic variants rs1218585 and rs1218584 are associated with spontaneous preterm birth in a Portuguese population

**DOI:** 10.1101/2025.05.11.25327390

**Authors:** Joana Couceiro, Carlos Família, Helena Marchão, Catarina Silva, José João Mendes, Pedro V. Baptista, Alexandra R. Fernandes, Alexandre Quintas

## Abstract

Spontaneous preterm birth (SPTB) is a major cause of neonatal morbidity and mortality worldwide. Although a genetic component has been implicated, specific variants remain poorly defined. The potassium calcium-activated channel subfamily N member 3 (*KCNN3*) gene encodes the SK3 potassium channel, which contributes to myometrial relaxation and uterine quiescence. This study aimed to assess the association between two *KCNN3* variants (rs1218585 and rs1218584) and SPTB in a Portuguese postpartum women cohort. Genotyping was performed using iPLEX Gold technology, and logistic regression was applied under multiple inheritance models, with adjustments made to sociodemographic and lifestyle variables. Both variants showed a significant association with SPTB. The GA genotype of rs1218585 and the CG genotype of rs1218584 were associated with 3.67- and 3.00-fold increased odds of SPTB, respectively. Associations remained significant after adjustment and multiple testing corrections. Haplotype analysis also identified a minor allele combination (AG) associated with increased odds of SPTB. These findings support a role for *KCNN3* variants in SPTB susceptibility and highlight the potential of SK3 potassium channels in preterm labour pathophysiology. Larger studies in diverse populations are warranted to validate these associations and clarify their potential clinical relevance.

## INTRODUCTION

Preterm birth (PTB), defined as birth before 37 completed weeks of gestation [1], is a major clinical and public health challenge [2]. Its estimated global prevalence is 9.9%, meaning that 1 in 10 babies was born preterm. PTB is the leading cause of death in children under five years old (18%) [3] and accounts for 70% of neonatal deaths and up to 75% of neonatal morbidity [4]. Beyond its health impact, PTB imposes substantial burdens on families and healthcare systems.

The aetiology of PTB remains unclear, but multiple risk factors have been identified, including socioeconomic factors and genetic predisposition, and about two-thirds of the cases are spontaneous (SPTB). Twin studies suggest that up to 40% of PTB cases have a genetic component [5,6] and familial aggregation studies further support a heritable influence. However, identifying specific genetic variants has been challenging. Despite numerous genetic association studies, many reported variants have not been consistently replicated or validated, highlighting the need for further research.

The potassium calcium-activated channel subfamily N member 3 (*KCNN3*), which encodes the small conductance calcium-activated potassium channel protein 3 (SK3), is among the candidate genes that remain poorly investigated in SPTB. SK3 channels are voltage-independent ion channels expressed in various tissues, including smooth muscle. In the myometrium, these channels play a crucial role in hyperpolarising the cell membrane, reducing contractile activity, thereby maintaining uterine quiescence during early gestation. As pregnancy progresses, SK3 expression is downregulated, increasing uterine excitability and facilitating labour onset [7–9]. Premature SK3 downregulation may lead to excessive uterine excitability and contractions, contributing to SPTB. Genetic variations in *KCNN3* have already been linked to PTB. Within the maternal genetic characteristics, Day et al. (2011) were the first to investigate *KCNN3* variants in SPTB, analysing 16 SNPs and identifying three – rs1218585 and rs1218584 among them – associated with this condition [10]. However, these two *KCNN3* variants have not been further explored. The growing recognition of SK3 channels in pregnancy and labour underscores the need for further investigation into their genetic regulation. As early as 2008, Pierce et al. suggested that targeting SK3 channels could help prevent or halt human preterm labour progression. In 2011, Day et al. emphasised the *KCNN3* gene as a logical candidate for SPTB research. However, few studies have explored the impact of *KCNN3* variants in this context. Thus, further research is needed to elucidate the role of this gene in SPTB and assess its potential as a biomarker. This study aimed to evaluate the association of the *KCNN3* variants rs1218585 and rs1218584 with SPTB in a Portuguese postpartum women cohort.

## MATERIALS AND METHODS

### Study Population

A case-control study was conducted involving 110 postpartum women recruited from the Puerperium Unit of the Gynaecology and Obstetrics Service at Hospital Garcia de Orta (HGO), Portugal, between March 2020 and May 2022. The case group included 32 women who experienced SPTB, defined as delivery before 37 completed weeks of gestation following spontaneous onset of labour. Exclusion criteria for cases included multiple pregnancies, congenital anomalies, and medically indicated PTB. The control group comprised 78 women with term deliveries (≥37 weeks). All participants provided fully informed written consent and completed a questionnaire addressing social-demographic and lifestyle characteristics. The study protocol was approved by the HGO Ethics Committee (No. 56/2019), and all procedures were conducted in accordance with the Declaration of Helsinki and the ethical principles for medical research involving human subjects. This study adheres to the Strengthening the Reporting of Genetic Association Studies (STREGA) guidelines, an extension of the STROBE statement [11].

### Sample collection and genotyping

A 2 mL peripheral blood sample was collected in EDTA-coated tubes from all enrolled participants (SPTB and controls) within 72 hours after delivery. Samples were anonymized by the hospital personnel using code identifiers, ensuring that the molecular laboratory technician was blinded to both patient information and group allocation.. The samples were immediately transported under refrigeration to the molecular laboratory at Egas Moniz School of Health & Science or the NOVA FCT Lab and stored at 4°C for a maximum of 24 hours before processing.

DNA was extracted using the NZY Tissue gDNA Isolation kit (NZYTECH), according to the manufacturer’s instructions, and stored at −80°C. Genotyping of two single nucleotide polymorphisms (SNPs) in the *KCNN3* gene (OMIM 602983) – rs1218585 (G>A) and rs1218584 (C>G) – was performed using the iPLEX^®^ Gold technology on the Agena Bioscience MassARRAY^®^ platform (CEGEN-FPGMX, Fundación Pública Galega de Medicina Xenómica, Santiago de Compostela, Spain), with a genotyping success rate above 98%.

### Statistical analysis

Data were analysed using R (v4.2.1) and PLINK (v1.9). Descriptive statistics were computed for all demographic and lifestyle variables. Comparisons between groups were conducted using chi-square or Fisher’s exact test for categorical variables and t-tests for continuous variables.

Hardy-Weinberg equilibrium (HWE) was tested in the control group. Minor allele frequency (MAF) for both SNPs were assessed and confirmed to exceed the commonly used 5% threshold prior to inclusion in model-based analyses.

The association between *KCNN3* variants and SPTB was initially assessed by estimating genotype frequencies and computing crude odds ratio (ORs) using contingency tables. The Wald test was used to determine the statistical significance of each genotype relative to a reference category. Power calculations and required sample sizes to achieve 80% power were estimated using OpenEpi. Subsequently, multiple genetic inheritance models were tested, including allelic, genotypic, dominant, recessive, and the Cochran-Armitage trend test (TREND, as a proxy for the additive model. Logistic regression models were then applied under the additive model to estimate crude and adjusted ORs. Adjustments were made for potential confounders identified in the literature, including age, ancestry, education level, occupation status, marital status, smoking habits, and dietary patterns. Corrections for multiple testing was applied using Bonferroni (B), Benjamini-Hochberg (BH), and Benjamini-Yekutieli (BY) methods. Additionally, linkage disequilibrium (LD) between the two variants, rs1218585 and rs1218584, was estimated using Haploview (v4.2). Haplotype frequencies were compared between groups, and their associations with SPTB were analysed using logistic regression. Empirical *p* values from 1,000 permutation tests were computed to validate haplotype associations.

## RESULTS

### Demographic and lifestyle information of the control group and PTB group

Table 1 presents the demographic and lifestyle data of women with SPTB (SPTB group) and women with full term pregnancy (control group). The study included 78 controls, aged 31.1 ± 5.71 years, and 32 women with SPTB, aged 32.0 ± 6.33 years. There was no significant difference in age between the two groups (p > 0.05). Regarding ancestry, most subjects in both groups self-identified as of European descent, with 87.2% in the control group and 84.4% in the SPTB group. No significant differences were found in other demographic parameters, including education level (p = 0.735), occupation status (p = 0.558), marital status (p = 0.610), and number of children (p = 0.787). Additionally, smoking habits did not differ significantly between the control and SPTB groups (p = 0.232), with most women in both groups being non-smokers (88.5% in controls and 78.1% in the SPTB groups).

**Table 1.** Demographic characteristics of mothers with SPTB and controls.

|  | Control<br>(N=78) | SPTB<br>(N=32) | *p* value |
| --- | --- | --- | --- |
| Age |  |  |  |
| Mean (SD) | 31.1 (5.71) | 32.0 (6.33) | 0.509 |
| Median [Min, Max] | 31.0 [22.0, 43.0] | 32.5 [18.0, 43.0] |  |
| Ancestry |  |  |  |
| European | 68 (87.2%) | 27 (84.4%) | 0.370 |
| African | 10 (12.8%) | 4 (12.5%) |  |
| Asian | 0 (0%) | 1 (3.1%) |  |
| Education level |  |  |  |
| Uneducated | 1 (1.3%) | 1 (3.1%) | 0.735 |
| Elementary school | 11 (14.1%) | 6 (18.8%) |  |
| High school | 35 (44.9%) | 14 (43.8%) |  |
| Graduate | 31 (39.7%) | 11 (34.4%) |  |
| Occupation status |  |  |  |
| Unemployed | 19 (24.4%) | 5 (15.6%) | 0.558 |
| Student | 2 (2.6%) | 1 (3.1%) |  |
| Employed | 57 (73.1%) | 26 (81.3%) |  |
| Marital status |  |  |  |
| Single | 32 (41.0%) | 11 (34.4%) | 0.610 |
| Cohabiting | 21 (26.9%) | 11 (34.4%) |  |
| Married | 23 (29.5%) | 8 (25.0%) |  |
| Divorced | 2 (2.6%) | 2 (6.3%) |  |
| Number of children |  |  |  |
| Mean (SD) | 1.50 (0.716) | 1.56 (1.22) | 0.787 |
| Median [Min, Max] | 1.00 [1.00, 4.00] | 1.00 [1.00, 7.00] |  |
| **Gestation age (weeks)** |  |  |  |
| <28 | 0 (0%) | 5 (15.6%) | <0.001* |
| 28-32 | 0 (0%) | 6 (18.8%) |  |
| 32-37 | 0 (0%) | 21 (65.6%) |  |
| ≥37 | 78 (100%) | 0 (0%) |  |
| **Previous PTB** |  |  |  |
| No | 74 (94.9%) | 25 (78.1%) | 0.013* |
| Yes | 4 (5.1%) | 7 (21.9%) |  |
| **Smoker** |  |  |  |
| No | 69 (88.5%) | 25 (78.1%) | 0.232 |
| Yes | 9 (11.5%) | 7 (21.9%) |  |
| **Dairy consumption** |  |  |  |
| Once a week | 1 (1.3%) | 3 (9.4%) | 0.075 |
| Once a day | 19 (24.4%) | 10 (31.3%) |  |
| Twice a day | 43 (55.1%) | 11 (34.4%) |  |
| Three times a day | 15 (19.2%) | 8 (25.0%) |  |
| **Cereals consumption** |  |  |  |
| Do not consume | 1 (1.3%) | 0 (0%) | 0.296 |
| Occasionally | 1 (1.3%) | 3 (9.4%) |  |
| Once a week | 6 (7.7%) | 0 (0%) |  |
| Once a day | 29 (37.2%) | 13 (40.6%) |  |
| Twice a day | 31 (39.7%) | 13 (40.6%) |  |
| Three times a day | 9 (11.5%) | 3 (9.4%) |  |
| Four times a day | 1 (1.3%) | 0 (0%) |  |

N – number of participants; SPTB – women with spontaneous preterm birth; SD – standard deviation. \* Parameter with statistically significant difference between control and PTB groups ( $p < 0.05$ ).

As expected, the gestation age at delivery was significantly lower in the SPTB group compared to the control group (p < 0.001). Among women with SPTB, 5 (15.6%) gave birth to extremely preterm babies (< 28 weeks of gestation), 6 (18.8%) had a very preterm birth (28 to 32 weeks), and 21 (65.6%) had a late PTB (32 to 37 weeks). Finally, consistent with the literature, the history of previous PTB was significantly more common in the SPTB group than in the control group (p = 0.013).

### Genotypes and haplotypes distributions in SPTB patients and controls

DNA from all participants was genotyped for the two selected SNPs, *KCNN3* rs1218585 and *KCNN3* rs1218584, to assess their potential associations with SPTB. Both SNPs fulfilled the HWE criteria (*p* > 0.05). Table 2 shows the significant genotype distributions between cases and controls, along with odds ratios (OR), corresponding 95% confidence intervals (CI), and respective power calculations. Both *KCNN3* variants, rs1218585 and rs1218584, were associated with significant differences in genotype distributions between SPTB patients and controls. For *KCNN3* rs1218585, the GA genotype was significantly associated with increased odds of SPTB compared to the GG reference genotype, with an OR of 3.67 (95% CI 1.54-8.70, *p* = 0.002). The rs1218585/AA genotype was not analysed, as it was absent in the SPTB group and observed in only one control participant. For *KCNN3* rs1218584, the CG genotype conferred a 3-fold increased odds of SPTB (OR = 3.00, 95% CI 1.27-7.10, *p* = 0.006). Again, the number of individuals homozygotes for the minor allele (GG) was too low to allow for a meaningful analysis (only one in the SPTB group and two in the control group). Thus, these results were excluded from the results presentation.

**Table 2.** Genotype distributions of *KCNN3* rs1218585 and rs1218584 in patients with SPTB and controls and power calculations.

| SNP | Genotype | Control | SPTB | OR | 95% CI | *p* value | Observed Power | Sample size<br>80% power |
| --- | --- | --- | --- | --- | --- | --- | --- | --- |
|  |  | N (%) | N (%) |  |  |  |  |  |
| *KCNN3* rs1218585 | GG | 57 (73.1) | 14 (43.8) | 1 (ref) |  |  |  |  |
|  | GA | 20 (25.6) | 18 (56.3) | 3.67 | 1.54 – 8.7 | 0.002 | 81.03% | NA |
| *KCNN3* rs1218584 | CC | 52 (66.7) | 13 (40.6) | 1 (ref) |  |  |  |  |
|  | CG | 24 (30.8) | 18 (56.3) | 3.00 | 1.27 – 7.10 | 0.006 | 62.3% | 140 |

CI – confidence interval; N – number of subjects; OR – odds ratio; SNP – single nucleotide polymorphism; SPTB – women with spontaneous preterm birth.

Table 3 summarises the chi-square statistics (χ^2^), degrees of freedom (DF), and p-values for the significant associations observed under the allelic model and the Cochran-Armitage trend test, which assumes an additive genetic effect. Both variants, rs1218585 and rs1218584, showed significant associations with SPTB in the allelic model (*p* = 0.014 and *p* = 0.030, respectively) and in the trend test, as tested by the Cochran–Armitage trend test (*p* = 0.007 and *p* = 0.021, respectively), supporting their involvement in susceptibility to SPTB.

**Table 3.** Association test results for *KCNN3* rs1218585 and rs1218584 under genetic inheritance models (allelic and trend tests)

| SNP | Test | $\chi^2$ | DF | *p* value |
| --- | --- | --- | --- | --- |
| *KCNN3* rs1218585 | TREND | 7.151 | 1 | 0.007 |
|  | ALLELIC | 5.998 | 1 | 0.014 |
| *KCNN3* rs1218584 | TREND | 5.344 | 1 | 0.021 |
|  | ALLELIC | 4.707 | 1 | 0.030 |

DF – degree of freedom; SNP – single nucleotide polymorphism; TREND – Cochran-Armitage trend test; $\chi^2$ – Chi-square test.

Table 4 presents the logistic regression analysis results, including the Wald test for crude (OR) and adjusted (aOR) odds ratios, with the latter accounting for potential confounders. This analysis confirmed that the *KCNN3* rs1218585 and rs1218584 variants were associated with increased odds of SPTB. For rs1218585, the crude OR was 2.97 (95% CI 1.31-6.76, *p =* 0.009). After adjusting for potential confounders, including age, ancestry, education level, occupation status, marital status, smoking status, and dairy and cereals consumption, the adjusted odds ratio (aOR) increased to 4.17 (95% CI 1.00-11.0, *p =* 0.004), remaining statistically significant after Bonferroni, BH, and BY corrections. For rs1218584, the crude OR was 2.37 (95% CI 1.38-4.12, *p =* 0.024). The adjusted OR was 2.86 (95% CI 1.23-6.70, *p =* 0.015), also remaining statistically significant after the same corrections (BH adjusted *p =* 0.023, BY adjusted *p =* 0.030, Bonferroni adjusted *p =* 0.045). These results indicate that both *KCNN3* variants contribute to SPTB, with the adjusted models further strengthening these associations.

**Table 4.** Crude and adjusted OR from logistic regression for the association of *KCNN3* rs1218585 and rs1218584 polymorphisms with SPTB.

| SNP | OR (95% CI) | *p* value | *p* value<sup>a</sup> | aOR (95% CI) | *p* value | *p* value<sup>a</sup> |
| --- | --- | --- | --- | --- | --- | --- |
| *KCNN3* rs1218585 | 2.97 (1.31-6.76) | 0.009 | 0.019(BH), 0.028(BY), 0.02(B) | 4.17 (1.58-11) | 0.004 | 0.008(BH), 0.012(BY), 0.008(B) |
| *KCNN3* rs1218584 | 2.37 (1.38-1.12) | 0.024 | 0.024(BH), 0.036(BY), 0.045(B) | 2.86 (1.23-6.7) | 0.015 | 0.015(BH), 0.023(BY), 0.030(B) |

aOR – odds ratio adjusted for age, ancestry, education level, occupation status, marital status, smoking status, and dairy and cereals consumption of the subjects; B – Bonferroni correction; BH – Benjamini-Hochberg correction; BY – Benjamini-Yekutieli correction; CI – confidence interval; OR – odds ratio; SNP – single nucleotide polymorphism; <sup>a</sup> – adjusted *p* value.

In addition, Figure 1 shows the LD analysis performed to assess the relationship between the two *KCNN3* variants. A haplotype analysis was also conducted to investigate further the potential role of rs1218585 and rs1218584 in SPTB susceptibility (Table 5).

**Table 5.** Haplotype frequencies of *KCNN3* rs1218585-rs1218584 in patients with SPTB and controls.

| Haplotype<br>(rs1218585-rs1218584) | Control (%) | SPTB (%) | OR | *p* value | Empirical *p*-value<sup>a</sup> |
| --- | --- | --- | --- | --- | --- |
| AG | 14.1 | 28.1 | 2.97 | 0.009 | 0.038 |
| GC | 82.1 | 68.8 | 0.42 | 0.024 | 0.046 |
| GG | 3.8 | 3.1 | 0.84 | 0.795<sup>†</sup> | NA |

OR – odds ratio; SPTB – spontaneous preterm birth; <sup>a</sup> – empirical *p*-value from permutation testing; <sup>†</sup> – non-significant association.

**Figure 1.**
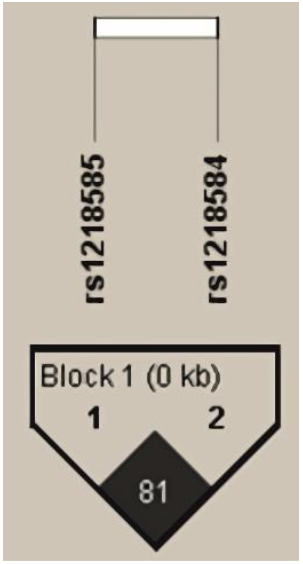
Linkage disequilibrium (LD) pattern among *KCNN3* variants. The two SNPs, rs1218585 and rs1218584, are in LD (r^2^ = 0.814), forming a haplotype block (Block 1)

Linkage disequilibrium analysis revealed that rs1218585 and rs1218584 were in strong LD (r^2^ = 0.814), forming a haplotype block (Figure 1). Haplotype analysis identified three possible haplotypes in the study population: AG, GC, and GG. The AG haplotype, formed by the minor alleles of both SNPs, was significantly more frequent in SPTB patients (28.1%) compared to controls (14.1%) and was associated with increased odds of SPTB (OR = 2.97, *p =* 0.009). The GC haplotype was more frequent in controls (82.1%) than in SPTB cases (68.8%) and was associated with lower odds of SPTB (OR = 0.42, *p =* 0.0242). the GG haplotype showed no significant association (*p >* 0.05). Permutation testing confirmed the significance of the AG haplotype (empirical *p =* 0.038) and the GC haplotype (empirical *p =* 0.046), suggesting that these *KCNN3* haplotypes may influence SPTB (Table 5).

## DISCUSSION

PTB remains a major global health concern [2], contributing to infant mortality [3,4], morbidity [4], and long-term developmental complications [12–18]. Genetic factors play a crucial role in susceptibility to PTB, particularly genes involved in inflammation and uterine physiology [19,20]. The present study investigated the association between two variants in the *KCNN3* gene (rs1218585 and rs1218584) and SPTB, providing further evidence of the involvement of *KCNN3* in pregnancy outcomes.

The main findings of this study were the association of *KCNN3* rs1218585 and rs1218584 with SPTB. Both SNPs showed a strongly significant association with this condition in all performed analysis. These findings are biologically supported by the established role of SK3 channels, encoded by *KCNN3*, in uterine physiology. In myometrial smooth muscle cells, potassium channels are crucial in maintaining gestation and triggering parturition [7–9,21], and their expression and density changes dynamically throughout pregnancy [9,22,23]. SK channels are constitutively associated with calmodulin, which regulates their activity by binding intracellular calcium, thus enabling channel opening [24]. Consequently, it allows for the voltage-independent transmembrane transfer of potassium ions across the cell membrane, generating a hyperpolarisation current that promotes smooth muscle relaxation and reduces contractile activity [25]. Alterations in SK expression or activity may impair proper repolarisation, potentially leading to aberrant uterine contractility [25]. The SK3 channels have a particularly significant role in gestation among SK family members. Studies in humans and mice [7–9,21] show that SK3 activity is paramount during pregnancy for uterine quiescence, maintaining pregnancy and prevent premature contractions [9,26]. From mid-to-late gestation, its downregulation increases uterine excitability, facilitating labour onset [7,9,22,27]. Consequently, SK3 overexpression may weaken uterine contractility, potentially delaying or preventing parturition [28], while premature SK3 dysfunction or reduced expression could enhance uterine excitability and increase SPTB risk [10]. Beyond uterine contractility, SK3 channels are involved in vascular remodelling during pregnancy, regulating placental blood flow and ensuring proper fetal development [29,30]. These findings highlight the multifaceted role of these channels in pregnancy, suggesting that dysregulations of SK3 activity may impact both uterine activity and gestational hemodynamics.

Genetic variants that modulate SK3 expression or function may influence uterine contractility and predispose to preterm labour. In a seminal study, Day and colleagues (2011) investigated 16 SNPs in the *KCNN3* gene and identified rs1218585 and rs1218584 associated with SPTB [10]. The authors showed that rs1218585 variant lie within a conserved intronic region, between exons 1 and 2, encompassing the promotor and an alternative exon (1c) that encodes a dominant-negative SK3 isoform, which suppresses channel activity and lowers cellular excitability [31]. However, these two variants have not been independently validated, and limitations in the original study – such as the lack of raw genotype data, limited statistical details, and absence of results after multiple testing corrections – warrant further investigation.

In the present study, we re-examined these SNPs in a case-control design and a more comprehensive statistical approach. Our findings reveal strong and statistically robust associations between both *KCNN3* rs1218585 and rs1218584 and SPTB in all the analyses performed, including within the logistic regression framework, even under the application of stringent significance corrections using the BH, BY, and Bonferroni methods.

Interestingly, no homozygous individuals for the minor allele of rs1218585 or rs1218584 were observed. According to data from the 1000 Genomes Project, the expected frequency of homozygotes for these SNPs is approximately 12% and 16%, respectively, in the overall population and about 23% for both SNPs in the European population. However, they were absent from our cohort of pregnant women. This finding raises the possibility that minor alleles in homozygosity could have a deleterious effect on pregnancy, potentially reducing the likelihood of carrying a pregnancy to term. Unfortunately, Day and colleagues do not provide raw genotype frequencies in their maternal DNA analysis, leaving it unclear whether they observed a similar pattern or if it is related to our small sample size. Although our study does not establish a direct causal link, the observed genotype distribution suggests that *KCNN3* polymorphisms may influence birth time onset. Further investigation using larger cohorts with longitudinal follow-up is needed to clarify potential effects on fetal survival and pregnancy outcomes.

LD analysis revealed a strong correlation between rs1218585 and rs1218584, suggesting the formation of a haplotype block. Notably, the haplotype composed of the minor alleles of both SNPs was also significantly associated with SPTB, further supporting the potential role of these *KCNN3* variants in pregnancy outcomes.

Finally, several studies have demonstrated that sociodemographic factors, namely maternal education [32– 35] and marital status [36,37], are associated with PTB. However, in our study, none of the analysed sociodemographic or lifestyle characteristics showed significant differences between cases and controls. To account for potential confounding, we performed logistic regression analyses adjusting for these variables, and the associations between *KCNN3* polymorphisms and SPTB remained statistically significant. These findings suggest that the observed genetic associations are independent of the evaluated demographic and lifestyle factors.

The main limitation of this study is the limited sample size (N = 110), which may affect the robustness of our findings and preclude further stratified analyses, such as assessing potential differences among SPTB subtypes (i.e. based on gestational age). This is the first independent study to replicate and support the association of rs1218585 and rs1218584 with SPTB. Despite the limited sample size, the observed associations are statistically robust and were obtained under strict methodological control, including multiple testing corrections and adjustment for confounders. These results are also biologically plausible, given the established role of SK3 channels in pregnancy. However, in line with recommendations for reporting genetic risk [38], further validation in larger and more diverse populations is essential to assess their potential translational relevance. The results of this study reinforce the role of *KCNN3* variants as potential contributors to the pathophysiology of preterm labour, which may open avenues to the development of early risk prediction tools, enabling clinicians to implement timely preventive strategies against PTB.

## Data Availability

All data produced in the present study are available upon reasonable request to the authors

## ACKNOWLEDGEMENTS

We are grateful to Professor Catarina Ramos and Professor Cristina Soeiro for their assistance with the development and analysis of the sociodemographic and lifestyle questionnaire. We also thank the nursing team of the Puerperium from the Service of Gynaecology and Obstetrics, Hospital Garcia de Orta, for their support in collecting blood samples from the participants. Additionally, we acknowledge Dr. Vanessa Machado and MSc. Patrícia Lyra for their valuable assistance in participant recruitment and questionnaire distribution.

This study is supported by Egas Moniz School of Health and Sciences (grant number: CEI/11/2018) and by national funds from FCT - Fundação para a Ciência e a Tecnologia, I.P., in the scope of the project UIDP/04378/2020 (10.54499/UIDP/04378/2020) and UIDB/04378/2020 (10.54499/UIDB/04378/2020) of the Research Unit on Applied Molecular Biosciences - UCIBIO and the project LA/P/0140/2020 of the Associate Laboratory Institute for Health and Bioeconomy - i4HB.

## STATEMENTS AND DECLARATIONS

### Compliance with Ethical Standards

This study was performed in line with the principles of the Declaration of Helsinki and adheres to the Strengthening the Reporting of Genetic Association Studies (STREGA) guidelines, an extension of the STROBE statement. Approval was granted by the Ethics Committee of Hospital Garcia de Orta, Almada, Portugal (Data 20.08.2019, No. 56/2019). Informed written consent was obtained from all individual participants included in the study.

### Statement of contribution

**Joana Couceiro**: investigation (lead); data curation; formal analysis (lead); writing – original draft preparation; final revision, visualization; project administration (supporting). **Carlos Família:** formal analysis (supporting); writing – review & editing. **Helena Marchal:** resources (supporting); investigation (samples collecting); project administration (hospital coordination). **Catarina Silva:** methodology (clinical diagnosis); project administration (hospital coordination); **José João Mendes:** funding acquisition; resources. **Pedro J Baptista:** conceptualization (supporting); validation; writing – review & editing. **Alexandra R Fernandes:** conceptualization (supporting); supervision; validation; project administration; funding acquisition; writing – review & editing. **Alexandre Quintas:** conceptualization (lead); supervision; validation; writing – original draft (supporting); writing – review & editing; visualization; funding acquisition; project administration. All authors read and approved the final manuscript.

